# Assessment and Prediction of Clinical Outcomes for ICU-Admitted Patients Diagnosed with Hepatitis: Integrating Sociodemographic and Comorbidity Data

**DOI:** 10.1101/2024.12.21.24319488

**Authors:** Dimple Sushma Alluri, Felix M. Pabon-Rodriguez

## Abstract

**Background:** Hepatitis, a disease characterized by inflammation of the liver, is a leading global health challenge that contributes to over 1.3 million deaths annually, with hepatitis B and C accounting for many of these fatalities. Intensive care unit (ICU) management of patients is particularly challenging due to the complex clinical care and resource demands. Despite advancements in ICU predictive analytics, limited research has specifically addressed hepatitis patients, creating a gap in optimizing care for this population.

**Methods:** This study focuses on predicting length of stay (LoS) and discharge outcomes and discharge location for ICU-admitted hepatitis patients using machine learning (ML) models. Leveraging data from the MIMIC-IV database, which includes around 94,500 ICU patient records, this study uses sociodemographic details, clinical characteristics, and resource utilization metrics to develop predictive models such as Random Forest, Logistic Regression, Gradient Boosting Machines, and Generalized Additive Model with Negative Binomial Regression.

**Results:** The ML models identified medications, procedures, comorbidities, age, and race as key predictors. Total LoS emerged as an important factor in predicting discharge outcomes and location.

**Conclusion:** This study demonstrates the value of machine learning models for predicting clinical outcomes for hepatitis patients, including length of stay and discharge status. The results underscore the influence of factors like race and age, revealing disparities that must be addressed in predictive care strategies. While the models show promise, challenges such as variability in prolonged stays and limited multi-class prediction accuracy point to the need for ongoing refinement and research.

**What is already known on this topic?:** Hepatitis B and C cause significant global mortality and often lead to critical illness requiring ICU care. ICU management for hepatitis patients is complex, and prolonged ICU stays are associated with increased costs and higher mortality risks. Although machine learning has been applied in ICU settings, few studies have focused on predictive modeling specifically for hepatitis patients.

**What this study adds?:** This study applies machine learning models to predict ICU length of stay, discharge outcomes, and discharge locations among hepatitis patients using a large, real-world dataset. It identifies key clinical and sociodemographic predictors, such as ICU medications, procedures, comorbidities, age, and race. The findings reveal that race is a consistent predictor across all models, highlighting underlying disparities in critical care outcomes.

**How might this study affect research, practice or policy?:** These predictive models can support more informed clinical decision-making, enhance ICU resource planning, and improve patient outcomes by identifying high-risk individuals early. The study also emphasize the need to address racial disparities in critical care outcomes through targeted research and policy changes.

## 1. INTRODUCTION

Hepatitis, an inflammatory liver disease, remains a significant global health challenge, claiming an increasing number of lives each year. According to the World Health Organization (WHO) 2024 Global Hepatitis Report, viral hepatitis is the second leading infectious cause of death globally, responsible for 1.3 million deaths annually (1). This number has risen from 1.1 million in 2019, with 83% of these deaths attributed to hepatitis B and 17% to hepatitis C (1). Every day, approximately 3,500 people die due to hepatitis B and C infections worldwide (2). Despite advancements in diagnostic tools and treatment options, testing and treatment coverage rates have plateaued, signaling a growing public health crisis (1). In the United States, the most common forms of viral hepatitis are hepatitis A, B, and C, each impacting the liver differently and predominantly affecting distinct populations ((3,4). Hepatitis B and C pose severe health risks, often leading to chronic conditions such as cirrhosis and liver cancer. These diseases are also the primary contributors to liver-related mortality globally (5). Managing intensive care unit (ICU) admitted patients with hepatitis is particularly challenging due to the treatment complexities and the resource-intensive nature of care required for severe cases.

One critical issue in ICU settings is the unpredictability of a patient’s length of stay (LoS), a key metric influencing hospital resource management, patient care quality, and healthcare efficiency. Prolonged ICU stays are closely linked to increased hospital costs and heightened resource strain (6). Research also indicates that longer ICU stays correlate with increased long-term mortality rates, underscoring the need for precise LoS predictions (7). Beyond LoS, discharge outcomes and locations serve as vital metrics for assessing patient recovery, readmission risks, and the burden on healthcare systems (8–11). Sociodemographic and clinical factors such as race, gender, marital status, insurance type, age, and type of hepatitis could significantly influence hospitalization outcomes, particularly for ICU-admitted hepatitis patients. These factors also play a crucial role in determining hospital stay duration and discharge outcomes (12–16). Recent years have seen the growing application of machine learning (ML) in healthcare, particularly predictive modeling. ML algorithms provide several advantages over traditional statistical approaches, including their ability to analyze large, complex datasets and identify subtle patterns that conventional techniques may overlook (17). Predictive models using ML have been successfully applied in ICU settings to improve forecasts related to patient outcomes, treatment responses, and resource needs (18,19). However, there is a noticeable lack of ML models tailored to hepatitis patients and their unique clinical profiles.

The objective of this study is to explore and compare ML models to predict the LoS, discharge location, and discharge outcomes for ICU-admitted hepatitis patients. These models will incorporate sociodemographic and clinical variables from ICU admission records as key predictors. By addressing this gap, the study aims to provide valuable insights for healthcare providers, enhancing resource planning and improving hepatitis patient outcomes in ICU settings.

## 2. MATERIALS AND METHODS

### 2.1. Data

The data set used for this study was sourced from the Medical Information Mart for Intensive Care (MIMIC-IV), version 3.0, a comprehensive and de-identified repository of patient records (20–22), spanning the years 2008- 2022. The MIMIC-IV comprises data collected from patients admitted to the emergency department (ED) or intensive care units (ICU) at the Beth Israel Deaconess Medical Center (BIDMC) in Boston, MA. It includes over 364,000 unique patient records, with a total of 546,028 hospital admissions and nearly 94,500 ICU stays, providing a rich source of information for developing predictive models in healthcare. MIMIC-IV is organized into two main data modules: *hosp* and *icu*. The *hosp* module captures information from the hospital-wide electronic health record (EHR), detailing hospitalizations, patient demographics, laboratory results, medication administration, billing data, and more. In contrast, the *icu* module contains highly detailed clinical data from the ICU, sourced from the MetaVision clinical information system, including treatment plans, and monitoring data (20–22). It is worth mentioning that this study was completed prior to the release of MIMIC-IV v3.1. We carefully considered all the notes from the release and determined that data extracted from v3.0 was appropriate for this analysis.

### 2.2. Variables

The covariates for this study include sociodemographic, clinical, and resource utilization data. Sociodemographic variables cover gender, age, race, marital status, and type of insurance. Admission characteristics include the type of admission and admission location. Clinical variables included the number of comorbidities, as well as indicators for Hepatitis A, B, C, D, and E, and Hepatic Coma. Resource utilization variables include counts of medications, procedures, and drugs dispensed before ICU admission. Additional ICU-specific measures capture the volume of fluids prescribed and the number of procedures performed within the ICU. This study aims to predict three primary clinical outcomes: *Total LoS*, measured as the total days spent in the ICU or hospital; *Discharge Location*, detailing the destination upon discharge, such as home, hospice, or skilled nursing; and *Discharge Outcome*, a binary variable indicating discharge status (dead/alive).

### 2.3. Data Processing and Preparation

Figure 1 illustrates the data pre-processing workflow for the study, detailing sequential steps taken to create the final dataset, completed using the data management system MySQL (23,24). To isolate hepatitis-related hospital admissions, we used the International Classification of Diseases (ICD) codes specific to viral hepatitis from a comprehensive table of diagnostic codes from the hospital billing information and linked to admission records. These were merged with demographic and length-of-stay data to create a hepatitis-specific cohort. We added pre- and post-ICU medication and procedure counts, created indicators for each hepatitis type to assess co-infections, and calculated comorbidity counts excluding hepatitis-related diagnoses. The combined dataset initially consisted of 4,409 patient records (Figure 1). After a data validation process, we identified 534 records with inconsistencies between discharge outcomes and discharge locations, reducing to a final dataset of 3,875 hepatitis patient records.

**Figure 1.**
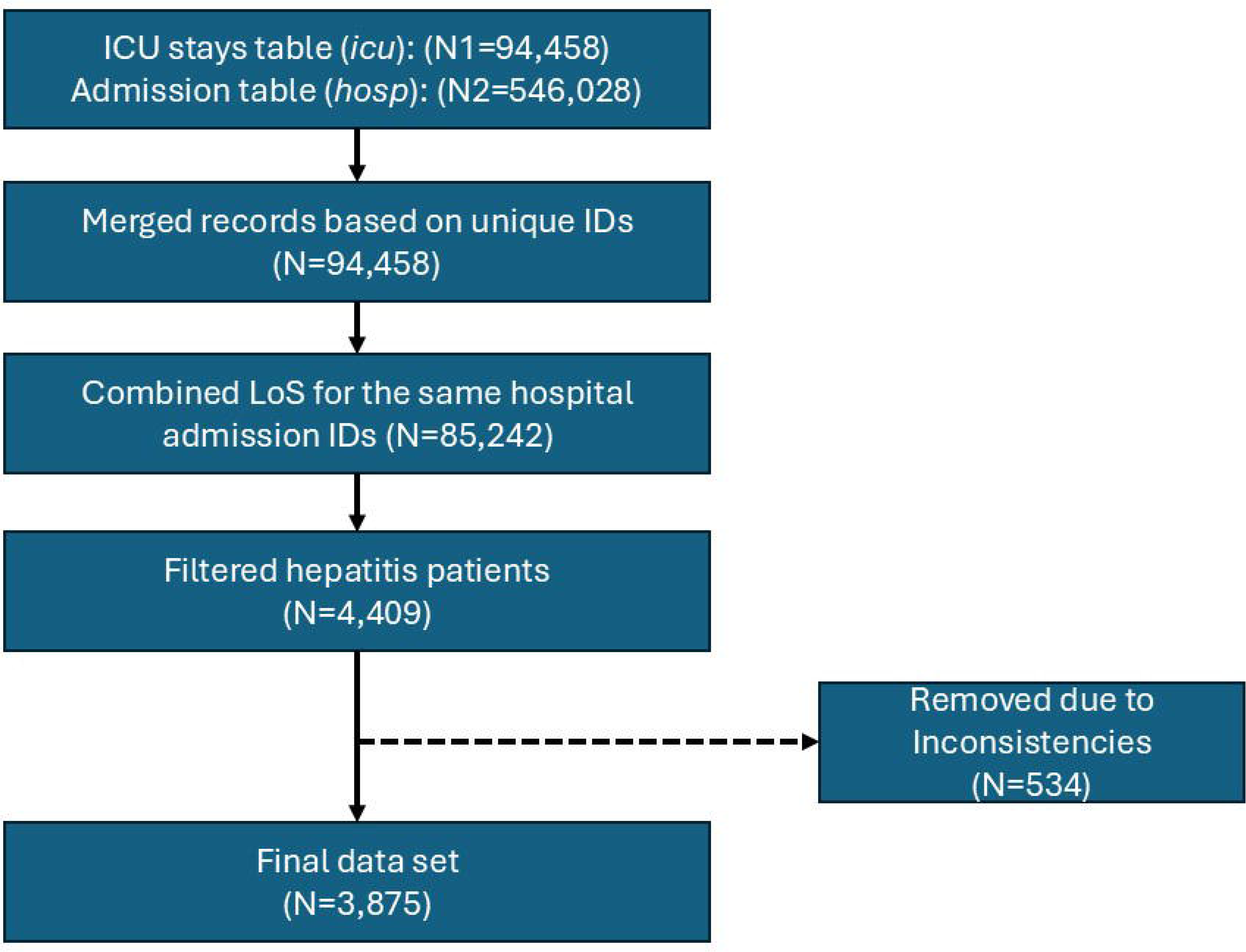
Data preprocessing workflow for extracting and refining hepatitis patient records from the MIMIC-IV database.

Missing data was handled via Multiple Imputation by Chained Equations (MICE) as appropriate (25). Predictive Mean Matching (PMM) was used for numeric columns, while Polytomous Logistic Regression (Polyreg) was applied to categorical variables with more than two levels. The discharge outcome variable displayed a significant class imbalance, with 3,353 records for the alive category and 522 for death. To address this, the Synthetic Minority Oversampling Technique (SMOTE) was used to balance the classes (26). Finally, an 80:20 split was used to separate the data into training and testing sets.

### 2.4. Machine Learning Models

For predicting the *discharge outcome*, a Logistic Regression (LR) and Random Forest Classification (RFC) models were used. Discharge location was not considered as a predictor in these models because certain discharge location categories overlapped with death records, providing redundant information. LR is a statistical model used for binary classification, estimating the probability of one of two possible outcomes (in this case alive or dead) based on a set of predictor variables (27). LR was chosen for its interpretability, providing clear insights into how each clinical and socio-demographic feature influences discharge outcome. On the other hand, RFC is applied to enhance the accuracy of predicting discharge outcome. The model constructs multiple decision trees using random data samples. The final prediction is determined by the majority vote across all trees. This ensemble approach captures complex, non-linear relationships among variables, offering strong predictive performance and minimizing the risk of overfitting (28). Although it does not offer the interpretive insights of LR, RFC’s capability to handle heterogeneous data makes it well-suited for maximizing predictive power in clinical scenarios (29).

To predict *LoS*, we considered two scenarios. In the first case, we considered LoS as a discrete (count) variable (30), using Generalized Additive Model (GAM) with a Negative Binomial (NB) distribution, denoted by GAM-NB, and as a continuous variable (31) in our second case via Random Forest Regression (RFR). For the discrete case, GAM-NB was utilized to address overdispersion (variance exceeds the mean) (32). By incorporating smooth functions, the GAM framework allows for modeling potential non-linear relationships between predictors and LoS (33). RFR, as another ensemble method, constructs multiple decision trees and averages their predictions to capture non-linear relationships and interactions among predictors (28).

For *discharge location*, a multi-class outcome (>2 levels), we compared a Gradient Boosting Model (GBM) and a Multinomial Logistic Regression (MLR). GBM is an ensemble learning technique that builds multiple decision trees sequentially, with each new tree correcting errors made by previous ones. This model focuses on reducing bias and improving predictive performance through boosting. GBM was chosen for its strong ability to handle complex, non-linear relationships and provide high accuracy (34). On the other hand, the MLR is an extension of logistic regression used when the dependent variable has more than two categories (35). It models the probability of each class as a function of the predictors. This model was included to establish baseline, offering a simpler and interpretable model for understanding the impact of predictor variables on different discharge locations. Supplemental Table 1 summarizes the models for each clinical outcome.

### 2.5. Model Evaluation and Resample

The models were evaluated using appropriate performance metrics (36) based on the type of outcome variable. For LoS, regression models were evaluated using Root Mean Squared Error (RMSE), R-squared (R²), and Mean Absolute Error (MAE). The multiclass classification models were assessed using Accuracy, the Kappa statistic, and the Brier Score. For the models based on binary outcome, we used Brier Score, Accuracy, Kappa, Area Under the Receiver Operating Characteristic Curve (ROC AUC), Sensitivity, and Specificity. Definitions for each of these performance metrics are given in the Supplemental Material. To ensure robustness and generalizability, a 10-fold cross-validation was employed (37), as it provides a balance between computational efficiency and reliability (37,38). K-fold cross-validation mitigate the risk of overfitting, provides more reliable estimates of the model’s performance, and enhances the reliability and generalizability of the results, making it a robust validation strategy for ML applications (38).

### 2.6. Software

Data analysis was conducted in R statistical software (39,40) using various R packages to streamline data manipulation, model development and predictions, and visualization (26,41–49). MySQL (23) was used to extract and manage data from the MIMIC-IV database. Statistical significance was assessed at the 5% significance level.

## 3. RESULTS

### 3.1. Demographic and Descriptive Statistics

The study included a total of 3,875 ICU-admitted patients with hepatitis-related conditions. The gender distribution was predominantly male, accounting for 68.2% (N = 2,644) of the sample, while females constituted 31.8% (N = 1,231). The average age of the participants was 53.26 years (SD = 12.77). The racial composition was primarily White (57.3%, N = 2,220), followed by Black (16.9%, N = 653), Hispanic/Latino (6.3%, N = 245), and Asian (6.0%, N = 234). Other racial groups, such as Indian/Alaska Native and Portuguese, made up a smaller proportion of the population, each representing less than 1% of the sample. These demographic characteristics, along with details on marital status, insurance type, clinical and admission characteristics, are summarized in Table 1. The distribution of hepatitis types is shown in Figure 2, which reveals that Hepatitis C was the most prevalent condition, affecting 82.2% (N = 3,186) of the total sample. Hepatitis B was the second most common condition at 19.3% (n = 746), while Hepatitis A (1.0%, N = 37), Hepatitis D (9.2%, N = 357), and Hepatitis E (<1%) were less frequent. Additionally, 8.0% (n = 311) of patients experienced hepatic coma. Most of the study participants were either single (48.3%, N=1,873) or married (28.5%, N=1,106). When looking at insurance type, a similar proportion of participants had either Medicaid (39.8%, N=1,542) or Medicare (38.0%, N=1,472). As expected, more than 50% of patients were admitted through the emergency ward (59.3%, N=2,296). The median number of comorbidities found was 18 (IQR: 12-24), while the number of medications (administered and dispensed) and procedures performed pre-ICU and during ICU, ranged (IQR) from 2 (pre-ICU procedures) to a high of 154 (ICU medications). The distribution of hepatitis types across racial groups is depicted in Figure 3. Hepatitis C was the most prevalent condition across all racial groups, with White patients contributing the largest proportion. Among Black and Hispanic/Latino patients, Hepatitis C was also the most common condition but showed slightly smaller proportions compared to White patients, while Hepatitis B was notably more prevalent among Asian patients compared to other racial groups. In addition, Hepatitis B disproportionately affects non-White individuals compared to Whites.

**Figure 2.**
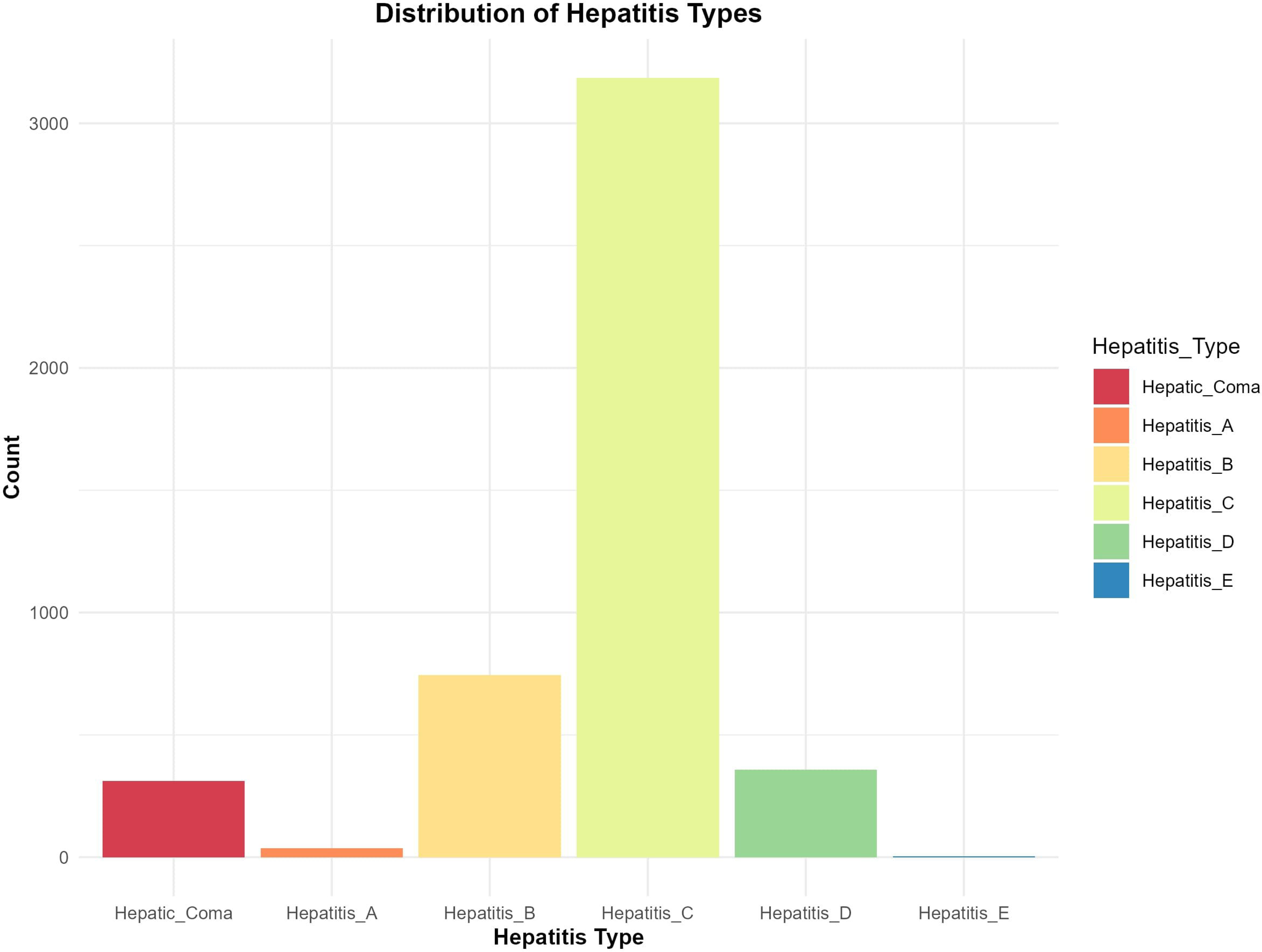
Distribution of hepatitis types among study participants.

**Figure 3.**
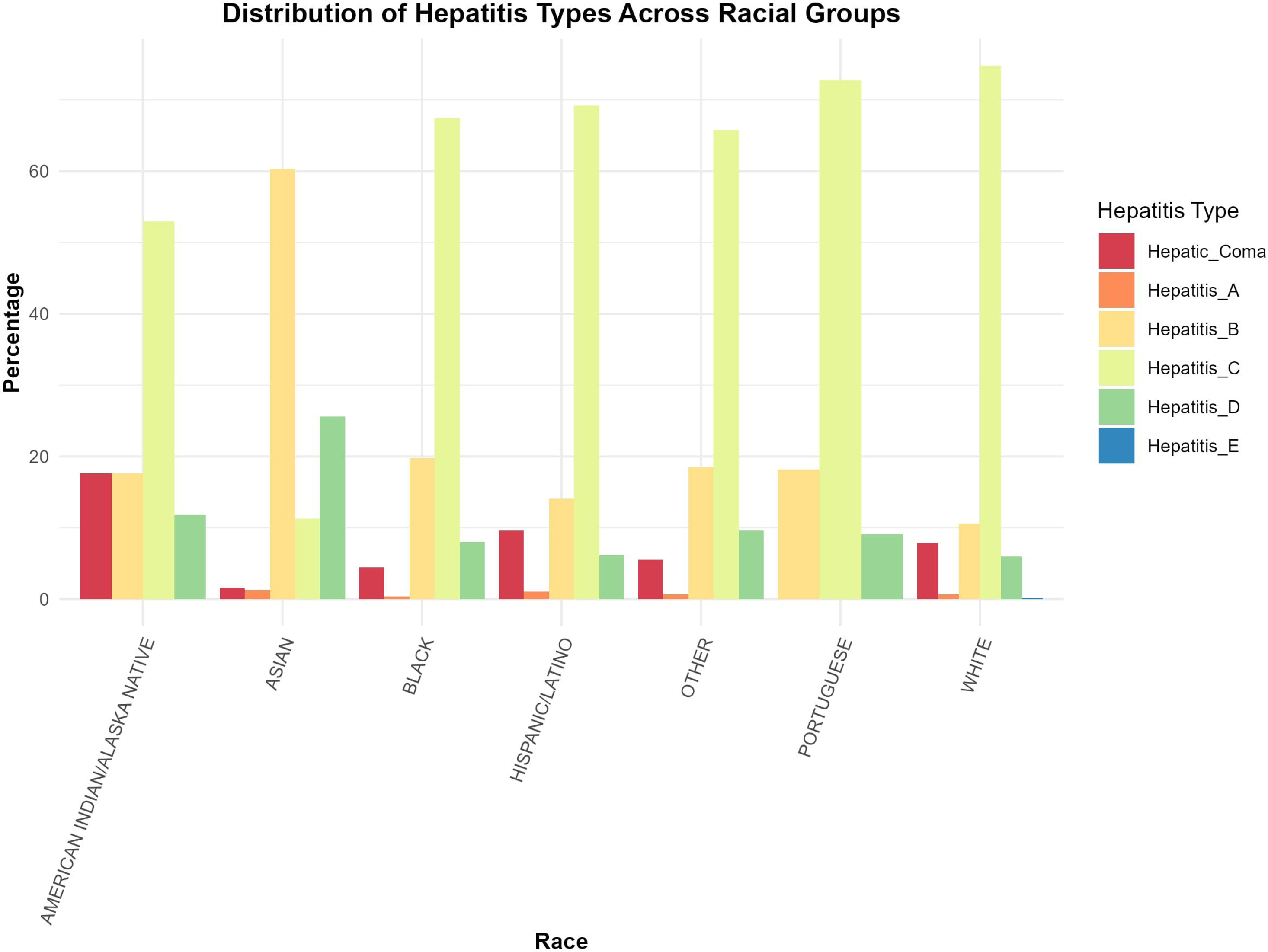
Distribution of hepatitis types across racial groups.

**Table 1.** Baseline demographic, clinical, and admission characteristics of study participants.

| Variable | N = 3,875 |
| --- | --- |
| Gender = Male (%) | 2644 (68.2) |
| Age (Mean (SD)) | 53.26 (12.77) |
| Race (%) |  |
| Indian/Alaska Native | 10 (0.3) |
| Asian | 234 (6.0) |
| Black | 653 (16.9) |
| Hispanic/Latino | 245 (6.3) |
| Other | 120 (3.1) |
| Portuguese | 29 (0.7) |
| Unknown | 364 (9.4) |
| White | 2220 (57.3) |
| Marital Status (%) |  |
| Divorced | 398 (10.3) |
| Married | 1106 (28.5) |
| Single | 1873 (48.3) |
| Widowed | 202 (5.2) |
| NA | 296 (7.6) |
| Insurance (%) |  |
| Medicaid | 1542 (39.8) |
| Medicare | 1472 (38.0) |
| Other | 123 (3.2) |
| Private | 674 (17.4) |
| NA | 296 (7.6) |
| Type Of Admission (%) |  |
| Direct Emergency | 230 (5.9) |
| Direct Observation | 10 (0.3) |
| Elective | 50 (1.3) |
| Emergency Unit Observation | 5 (0.1) |
| Emergency Ward | 2296 (59.3) |
| Observation Admit | 548 (14.1) |
| Surgical Same Day Admission | 155 (4.0) |
| Urgent | 581 (15.0) |
| Number of Comorbidities (Median [IQR]) | 18.00 [12.00, 24.00] |
| Pre-ICU Medications Count (Median [IQR]) | 82.00 [36.00, 140.25] |
| Pre-ICU Procedures Count (Median [IQR]) | 3.00 [2.00, 6.00] |
| Dispensed Medications Count (Median [IQR]) | 65.50 [33.00, 118.50] |
| ICU Medications Count (Median [IQR]) | 54.00 [20.00, 154.00] |
| ICU Procedures Count (Median [IQR]) | 7.00 [3.00, 14.00] |

The distribution of LoS for hepatitis patients was highly skewed as commonly seen in hospital records, with most patients having a short hospital stay as shown in Supplemental Figure 1. The median total LoS was 2 days (IQR: 1–4), with some patients having an extended stay of over 30 days. Variations in LoS were also observed based on admission location (Supplemental Figure 2). Patients admitted from emergency admissions had shorter median stays compared to those transferred from other facilities. This figure highlights how admission pathways may influence the duration of ICU stays. Supplemental Figure 3 illustrates the relationship between the total LoS and discharge outcome. Patients with longer LoS were more likely to have died, as indicated by the higher median and wider range of LoS in the death group compared to the alive group. Supplemental Figure 4 on the other hand, highlights the distribution of discharge locations among patients. The majority (51.5%, n = 1,997) were discharged to home-based care. Skilled nursing or long-term care facilities accounted for 17.7% (n = 684). Other notable discharge destinations included rehabilitation centers, hospice care, psychiatric care, other healthcare facilities, and patients leaving the hospital against medical advice, each contributing less than 5% of the sample.

### 3.2. Model Results and Performance

#### Discharge Outcome (Dead/Alive)

The LR and RFC models were developed and evaluated to predict discharge outcomes. A complete set of performance metrics for both models are presented in Supplemental Table 2. On the test data, LR yielded a Brier Score of 0.1302, an accuracy of 0.818, a Kappa of 0.622, and an ROC AUC of 0.887. Sensitivity and specificity were 0.73 and 0.87, respectively. The RFC model demonstrated a lower Brier Score of 0.0875, an accuracy of 0.87, a Kappa of 0.742, and a higher ROC AUC of 0.95. Sensitivity and specificity for the Random Forest model were 0.78 and 0.94, respectively. Cross-validation results are also shown in Supplemental Table 2, indicating the mean and standard deviation for each metric across repeated samples. The mean Brier Score for RFC was 0 089 (±0.010), while LR had a mean Brier Score of 0.129 (±0.011). The RFC model had a mean accuracy of 0.87 (±0.017) and a mean Kappa of 0.729 (±0.037), compared to LR’s mean accuracy of 0.82 (±0.02) and mean Kappa of 0.629 (±0.043).

Statistical comparisons were conducted to evaluate differences in performance metrics between the LR and RFC models. The results indicated statistically significant differences (p < 0.001) for all key metrics, including Brier Score, accuracy, Kappa, ROC AUC, sensitivity, and specificity. The RFC model consistently outperformed LR across all evaluated criteria for predicting discharge outcomes for the ICU-admitted hepatitis patients. Figures 4 and 5 display the variable importance for the LR and RFC models, respectively. The top predictors in the LR model (Figure 4) were pre-ICU medications count, ICU medications count, and ICU procedures count, with sociodemographic factors such as race and age also ranking among the top 10 variables. For the RFC model (Figure 5), ICU medications count, ICU procedures count, and total LoS were identified as the most influential variables, with race and age again appearing among the top 10 predictors. Supplemental Table 3 summarizes the confusion matrix results for both models. Both models showed good sensitivity and specificity values, with RFC performing better with higher precision.

**Figure 4.**
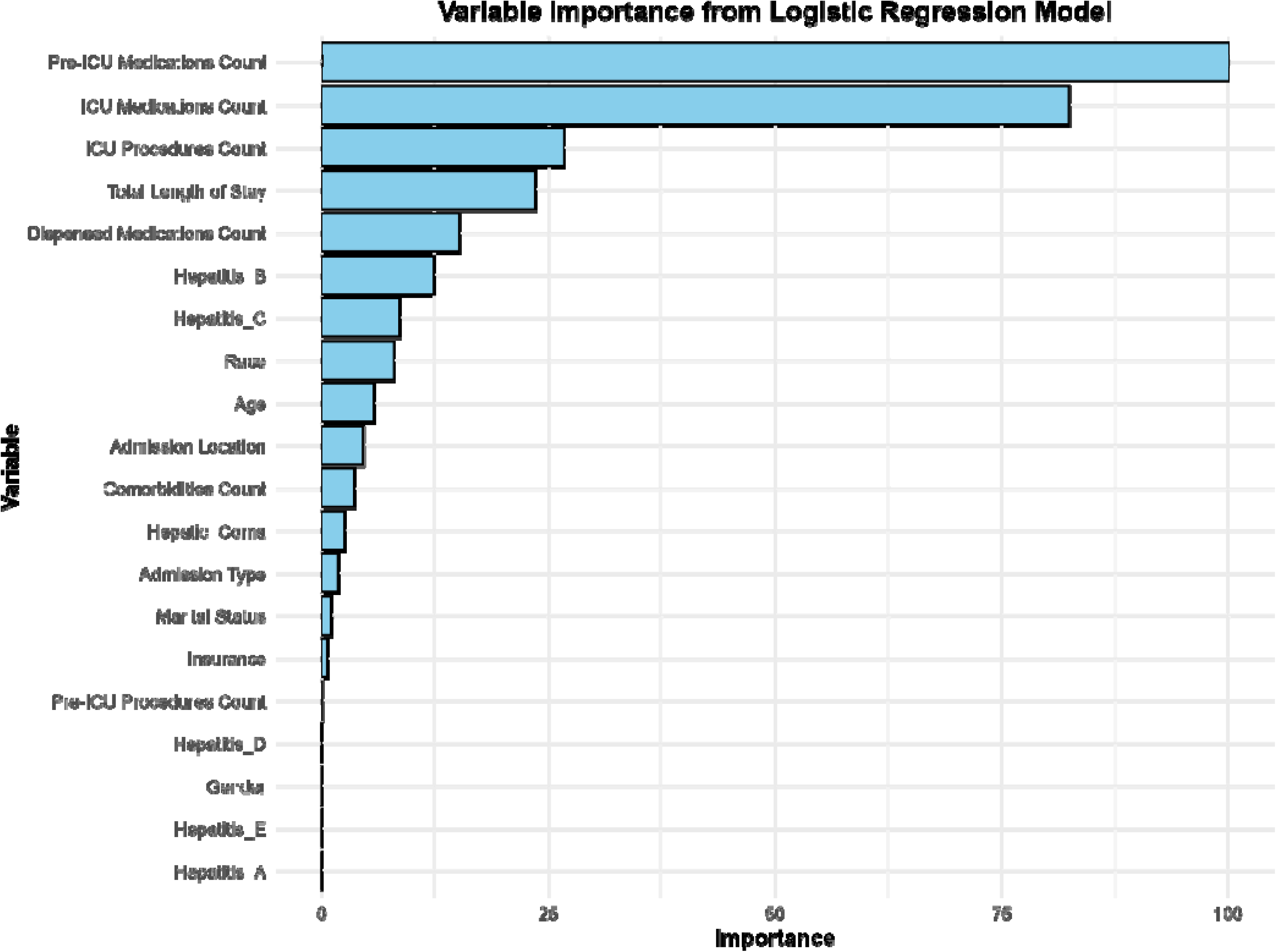
Variable Importance for Logistic Regression for Discharge Outcome.

**Figure 5.**
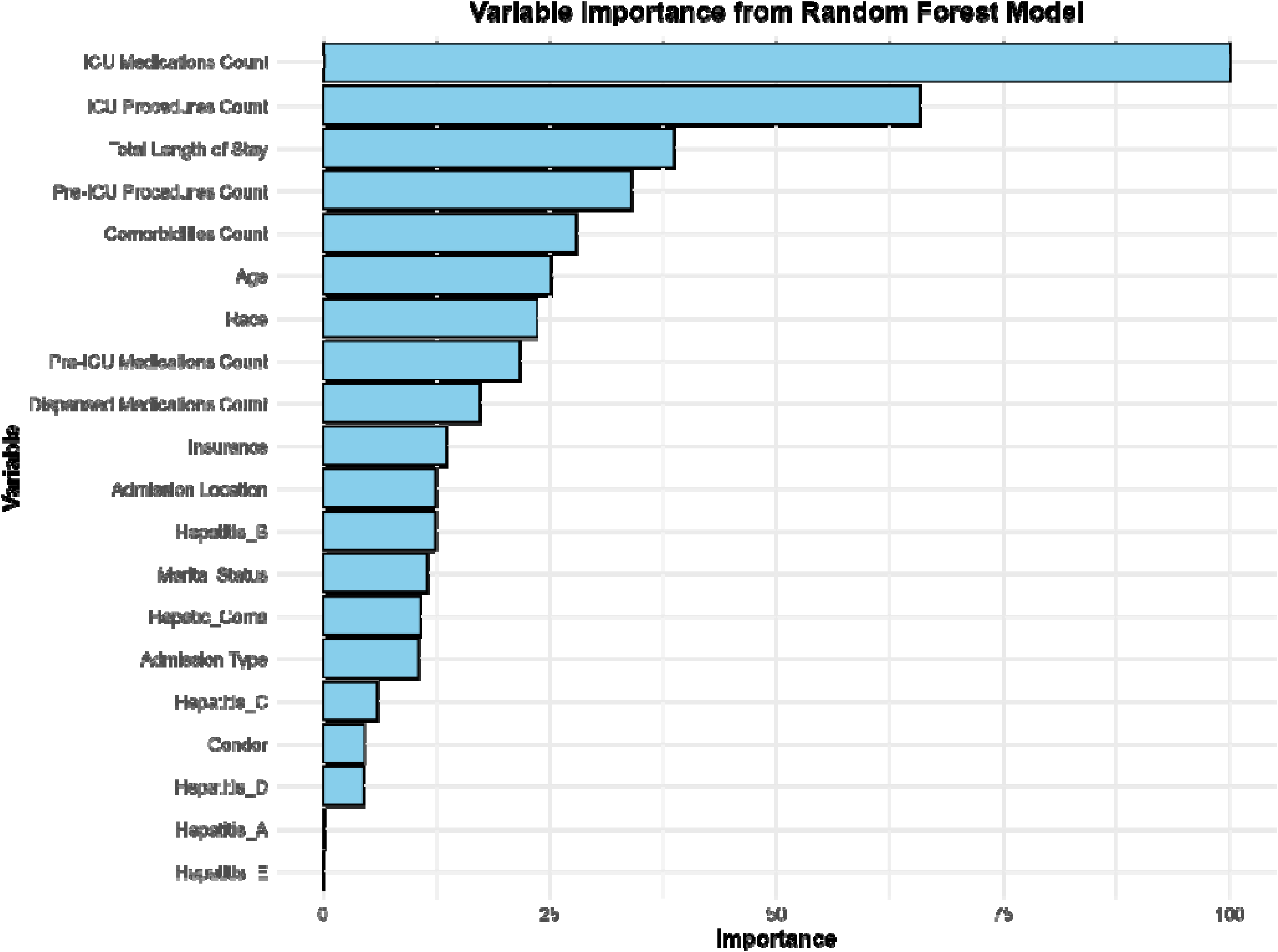
Variable Importance for Random Forest Classification for Discharge Outcome.

#### Total Length of Stay

The GAM-NB and RFR models were used to predict total LoS as discrete and continuous outcome. The mean LoS in the dataset was 3.69, with a variance of 42.54, indicating overdispersion. On the test data, the GAM-NB model achieved a RMSE of 2.9619, a MAE of 1.4237, and an R^2^ of 0.7594, demonstrating its ability to explain 75.94% of the variance (Supplemental Table 4). The cross-validated estimates are RMSE of 5.5212, mean R^2^ of 0.7601, and mean MAE of 1.5634. As shown in Supplemental Figure 5, most predictions are closely aligned with actual values, deviations are observed at higher LoS, indicating potential challenges in predicting extended stays. The RFR model, applied after log-transforming the total LoS, resulted in improved performance, achieving an RMSE of 0.3144, an R^2^ of 0.821, and an MAE of 0.2377 on the test data. Cross-validation further confirmed the model’s consistency, with mean RMSE of 0.295 (±0.020), mean R^2^ of 0.838 (±0.021), and mean MAE of 0.229 (±0.014). However, when done in the original scale, the performance metrics were: RMSE of 3.271, R^2^ of 0.755, and MAE of 1.360 (Supplemental Table 5).

Figure 6 illustrates the variable importance of the RFR model for predicting LoS. The most influential predictors were the number of ICU medications and procedures, comorbidities count, and pre-ICU procedure count. While clinical factors were most prominent, sociodemographic variables such as age and race also emerged as important predictors. Supplemental Figure 6 illustrates the relationship between predicted and actual LoS values for the RFR model. The plot on the left presents the scatter plot for the log-transformed data, where a strong linear relationship is observed, with predictions closely aligning with actual values. In contrast, the plot on the right displays greater dispersion for higher values.

**Figure 6.**
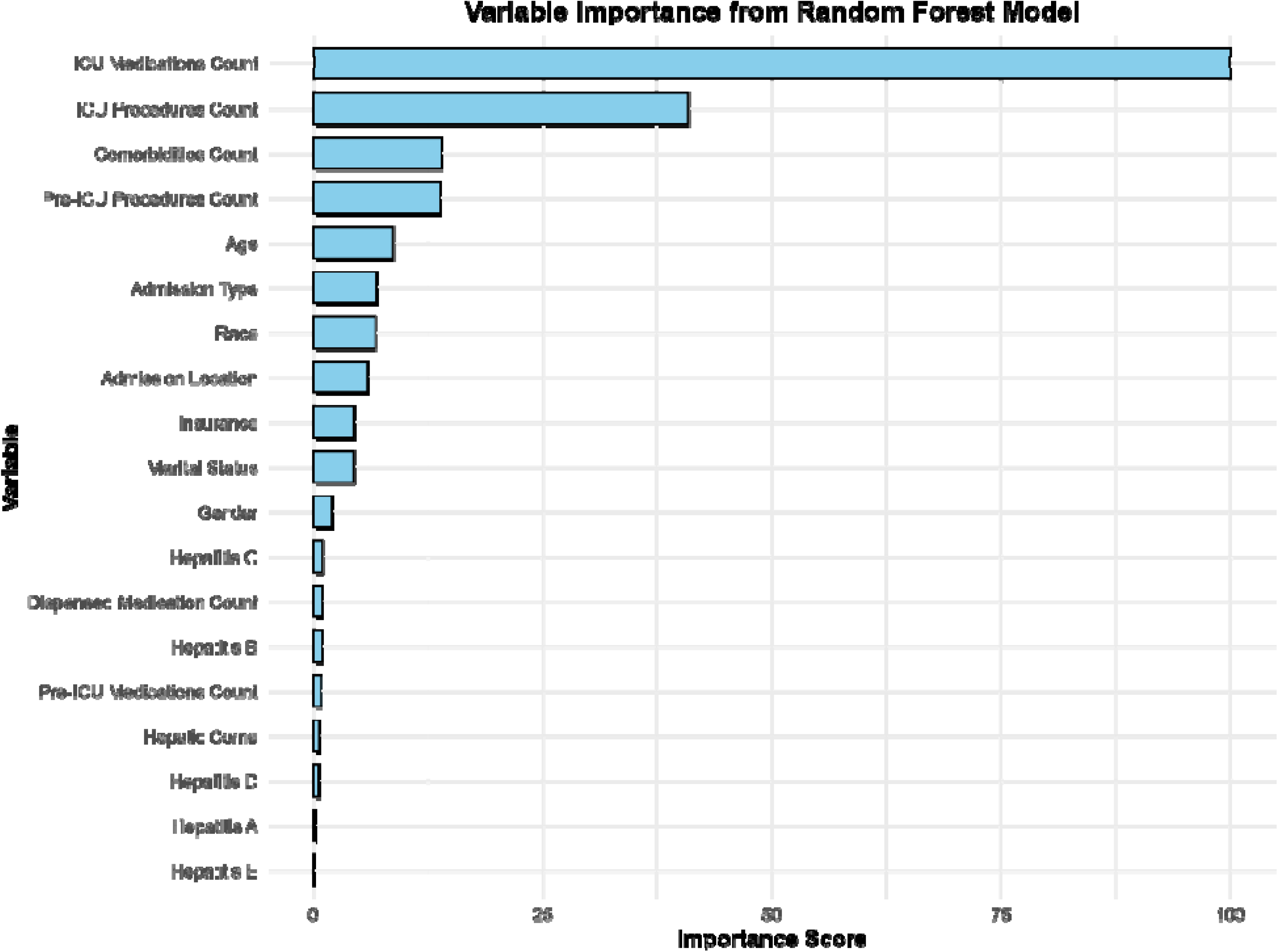
Variable Importance for Random Forest Regression Model for Total LoS.

#### Discharge Location

The GBM was evaluated on the test data, achieving a Brier Score of 0.589, an accuracy of 0.558, a Kappa of 0.211, and an ROC AUC of 0.7407 (Supplemental Table 6). Cross-validation analysis resulted in mean Brier Score of 0.595 (±0.0206), mean accuracy of 0.559 (±0.227), mean Kappa of 0.2077 (±0.0391), and mean ROC AUC of 0.7319 (±0.0282), indicating stable performance across folds. The MLR produced an accuracy of 0.5639 on the test data, with a Kappa value of 0.2037, and Brier Score of 0.5896 (Supplemental Table 6). Cross-validation yielded consistent results: mean accuracy of 0.5658 (±0.15), mean Kappa of 0.2335 (±0.30), mean Brier Score of 0.6 (±0.0352), suggesting comparable predictive performance (Supplemental Table 6). Significant predictors identified in this model included gender, marital status, insurance type, type of admission, number of comorbidities, and number of ICU procedures and medications. Figure 7 illustrates the variable importance of GBM, highlighting the number of ICU medications, comorbidities, age, total LoS, and race as the most influential predictors.

**Figure 7.**
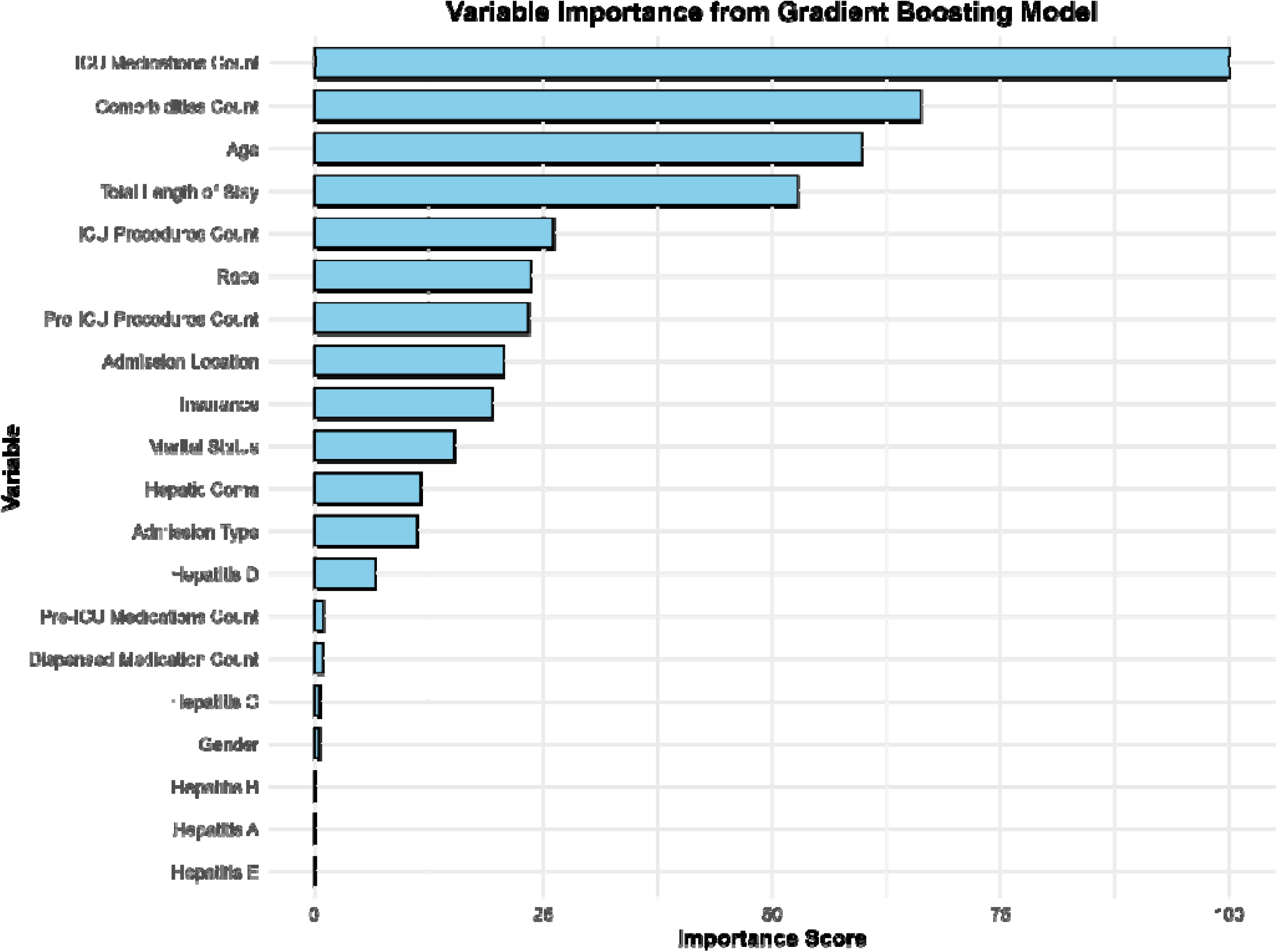
Variable Importance for Gradient Boost Model for Discharge Location.

## 4. DISCUSSION

Despite significant advancements in critical care for hepatitis-related conditions, this study highlights the complexities associated with ICU LoS, discharge outcomes, and discharge locations. By leveraging ML models, we predicted these outcomes and identified key predictors, including ICU procedures, medication counts, comorbidities, and sociodemographic factors such as age and race. Notably, race consistently emerged as one of the top predictors across all models, underscoring its critical role in influencing health outcomes for hepatitis patients. This finding is particularly significant as it aligns with observed racial disparities in the raw data, suggesting systemic inequities. Additionally, Hepatitis C emerged as the most prevalent condition in our cohort, aligning with global epidemiological trends that underscore its significant burden on critical care systems (50,51).

While predicting LoS, RFR model demonstrated superior predictive accuracy, particularly after applying a log transformation to stabilize variance. The log-transformed RFR achieved an R^2^ of 0.82 and an RMSE of 0.31 on test data, effectively capturing complex interactions among predictors. By comparison, GAM-NB achieved an R² of 0.76 and RMSE of 2.96, effectively addressing overdispersion but yielding higher errors. The key predictors for LoS included ICU medication counts, ICU procedure counts, comorbidities, pre-ICU procedures, and patient age. These findings align with prior studies (52), which demonstrated that higher procedural and pharmacological interventions often correlate with disease severity and resource intensity, leading to longer ICU stays. Similarly, another study by Tola Getachew Bekele et al. (53) emphasized that prolonged ICU stays are associated with complications, readmissions, and sedative use. Although the models performed well for shorter stays, predicting longer LoS posed challenges due to increased variability and extreme values, as reflected in Supplemental Figures 5 and 6. In a study by Xu et al. (31), the authors observed similar challenges, reporting that ML models often struggle to predict longer stays due to the inherent variability of extended ICU durations. Our study reflects these findings, with greater error dispersion observed in the upper range of LoS predictions. The performance of RFR in predicting LoS aligns with Iwase et al. (54), who reported high predictive accuracy for ICU stays using RFR. Ensemble methods such as RFR excelled in capturing nonlinear interactions and handling heterogeneous predictors, as further evidenced by the work of Alghatani et al. (55), who achieved 65% accuracy for LoS predictions in ICU datasets.

The prediction of discharge outcomes was conducted using LR and RFC models. RFC outperformed LR across all metrics, achieving an accuracy of 87.5% and a ROC AUC of 0.95 on test data, compared to the metrics by LR, achieving an accuracy of 81.67% and an ROC AUC of 0.89. Statistically significant predictors for discharge outcomes included ICU medication counts, ICU procedure counts, and total LoS, all of which reflect disease severity and the intensity of care and key variables in predicting LoS. Prolonged LoS has been linked to increased mortality, consistent with Moitra et al. (7), who found that each additional day in the ICU beyond seven days significantly raises mortality risk. Similar findings were reported by Lingsma et al. (56) and Tola Getachew Bekele et al. (53), who highlighted the correlation between extended LoS and mortality. The sociodemographic factors also played a crucial role in predicting discharge outcomes. Race, for instance, emerged as a significant predictor in our study, aligning with Olanipekun et al. (57), who found disparities in ICU mortality among demographic groups.

The prediction of discharge location presented unique challenges, primarily due to the small number of records and large number of categories (total of 8). GBM and MLR yielded moderate performance for this multi-categorical outcome, with accuracies of 55.87% and 56.39%, respectively. These results highlight the complexities of multi-class prediction in critical care settings with limited data. SMOTE was applied to address class imbalance but resulted in data loss, which limited the model’s performance. Abad et al. (58) addressed similar challenges using hierarchical classifiers and SMOTE to manage imbalanced multi-class datasets, suggesting that advanced techniques and larger datasets are critical for improving predictive accuracy. Additionally, the authors noted that discharge decisions are influenced by subjective factors such as caregiver preferences, resource availability, and social support, which may not be fully captured by clinical and demographic data alone. Top predictors for discharge location included gender, marital status, insurance type, admission type, number of comorbidities, and ICU procedures and medications. These findings align with Mickle and Deb (59), who highlighted the importance of clinical and patient characteristics in discharge planning. The association of ICU procedures and medications with discharge location reflects the intensity of care required, further emphasizing their relevance in critical care contexts.

This study had several limitations that warrant further consideration. First, the reliance on the MIMIC-IV database, which reflects a single institution’s patient population and care practices, may limit the generalization of the findings to other healthcare settings with differing demographics and protocols. Second, the imbalanced distribution of outcomes, particularly for rare discharge destinations (e.g., hospice, against medical advice), may have reduced model stability and predictive accuracy. While ML models like RF demonstrated strong performance, their limited interpretability compared to simpler models like LR poses challenges for clinical adoption. Future research should focus on validating these findings across diverse datasets to enhance generalizability. Addressing outcome imbalances through advanced techniques without data loss will be critical for better prediction of rare outcomes like hospice discharge or leaving against medical advice. Moreover, future studies could explore fitting a GAM with a Zero-Inflated Negative Binomial (ZINB) distribution to account for overdispersion and zero inflation simultaneously. Expanding this work to include real-time predictive tools integrated into Clinical Decision Support Systems (CDSS) could provide dynamic insights for resource allocation and care optimization. Finally, although not a limitation, it would be recommended and appropriate to consider the most recent version of the MIMIC-IV database to account for all updates and changes. As mentioned in the Methods section, this study was completed prior to the release of v3.1. We carefully considered all the notes from the release and determined that data extracted from v3.0 was appropriate for the analysis.

## 5. CONCLUSION

This study highlights the potential of ML in advancing critical care for hepatitis patients. The models identified key predictors of ICU LoS, discharge outcomes, and discharge locations, with clinical factors such as pre-ICU and ICU procedures and medication counts playing a significant role in determining these outcomes. Additionally, sociodemographic factors, including age and race, were consistently identified as important predictors, highlighting the presence of racial disparities among ICU-admitted patients diagnosed with hepatitis. These findings highlight the importance of addressing such disparities while using predictive analytics to optimize resource allocation, improve patient outcomes, and guide targeted interventions. Despite its strengths, the study also revealed challenges such as variability in extended LoS and limited data for multi-class predictions, underscoring the need for further advancements in predictive modeling strategies in these areas. Continued research in this area is essential not only to enhance predictive models but also to support broader public health efforts aimed at preventing, detecting, and managing hepatitis, ultimately reducing the burden of liver disease and hepatitis-related complications.

## 8. ETHICS STATEMENT

The Indiana University Human Research Protection Program (HRPP) staff determined the analysis done in this study was not human subject research and did not require further Institutional Review Board (IRB) review before conducting the study with MIMIC-IV v3.0. The study was conducted under the data use agreement governing MIMIC-IV, ensuring compliance with ethical standards for research involving human subjects.

## 9. DATA AVAILABILITY STATEMENT

The datasets used for this study are available at <u>MIMIC-IV v3.0</u> only for credentialed users following the PhysioNet Credentialed Health Data Use Agreement (DUA).

