## Supplemental Material for "Assessment and Prediction of Clinical Outcomes for ICU-Admitted Patients Diagnosed with Hepatitis: Integrating Sociodemographic and Comorbidity Data"

Sup Table 1. Machine learning models chosen for predicting each of the health outcomes for hepatitis patients.

| **Outcome** | **Type** | **Model** |
| --- | --- | --- |
| Discharge Outcome (Death/Alive) | Binary | Logistic Regression (LR) |
|  |  | Random Forest Classification (RFC) |
| ICU Length of Stay (LoS) | Count | Generalized Additive Model (GAM) |
|  | Continuous | Random Forest Regression (RFR) |
| Discharge Location | Categorical (>2 levels) | Gradient Boosting Model (GBM) |
|  |  | Multinomial Logistic Regression (MLR) |

*Sup Table 2. Performance metrics of Logistic Regression and Random Forest models on test and cross-validated data.*

| **Metrics** | **Logistic Regression** | | **Random Forest** | |
| --- | --- | --- | --- | --- |
|  | **Test data** | **Cross-validation** | **Test data** | **Cross-validation** |
| Brier Score | 0.1301 | 0.129 ± 0.011 | 0.0877 | 0.089 ± 0.010 |
| Accuracy | 0.8167 | 0.821 ± 0.020 | 0.8755 | 0.870 ± 0.017 |
| Kappa | 0.6220 | 0.629 ± 0.043 | 0.7421 | 0.729 ± 0.037 |
| ROC AUC | 0.8865 | 0.889 ± 0.017 | 0.9499 | 0.948 ± 0.012 |
| Sensitivity | 0.7350 | 0.736 ± 0.040 | 0.7823 | 0.780 ± 0.033 |
| Specificity | 0.8792 | 0.884 ± 0.019 | 0.9469 | 0.937 ± 0.016 |

Sup Table 3. Confusion matrix metrics for Logistic Regression and Random Forest models

| **Metric** | **Logistic Regression** | | **Random Forest** | |
| --- | --- | --- | --- | --- |
|  | **Class 0** | **Class 1** | **Class 0** | **Class 1** |
| Agreement | 0.4431 | 0.2975 | 0.4836 | 0.3261 |
| Sensitivity | 0.7737 | 0.6962 | 0.8444 | 0.7632 |
| Specificity | 0.6962 | 0.7737 | 0.7632 | 0.8444 |
| PPV (Precision) | 0.7734 | 0.6966 | 0.8270 | 0.7854 |
| NPV | 0.6966 | 0.7734 | 0.7854 | 0.8270 |

Sup Table 4. Performance metrics of General Additive Model with Negative Binomial

| **Metric** | **Test Data** | **Cross-Validation** |
| --- | --- | --- |
| Root Mean Squared Error | 2.9619 | 5.5212 |
| Mean Absolute Error | 1.4237 | 1.5634 |
| R-squared | 0.7594 | 0.7601 |

Sup Table 5. Performance Metrics of Random Forest Regression Models

| **Metric** | **Test Data** | **Log Transformed Test Data** | **Cross-Validation** |
| --- | --- | --- | --- |
| Root Mean Squared Error | 3.271 | 0.3144 | 0.295 (±0.020) |
| Mean Absolute Error | 1.360 | 0.2377 | 0.229 (±0.014) |
| R-squared | 0.755 | 0.821 | 0.838 (±0.021) |

Sup Table 6. Performance metrics of Gradient Boosting and Multinominal Regression models on test and cross-validated data.

| **Metric** | **Gradient Boosting Model** | | **Multinominal Regression** | |
| --- | --- | --- | --- | --- |
|  | **Test Data** | **Cross Validation** | **Test Data** | **Cross Validation** |
| Accuracy | 0.5587 | 0.5591 (±0.2270) | 0.5639 | 0.5658 (±0.15) |
| Kappa | 0.2110 | 0.2077 (±0.0391) | 0.2037 | 0.2335 (±0.30) |
| Brier Score | 0.5898 | 0.5950 (±0.0206) | 0.5896 | 0.6 (±0.0352) |

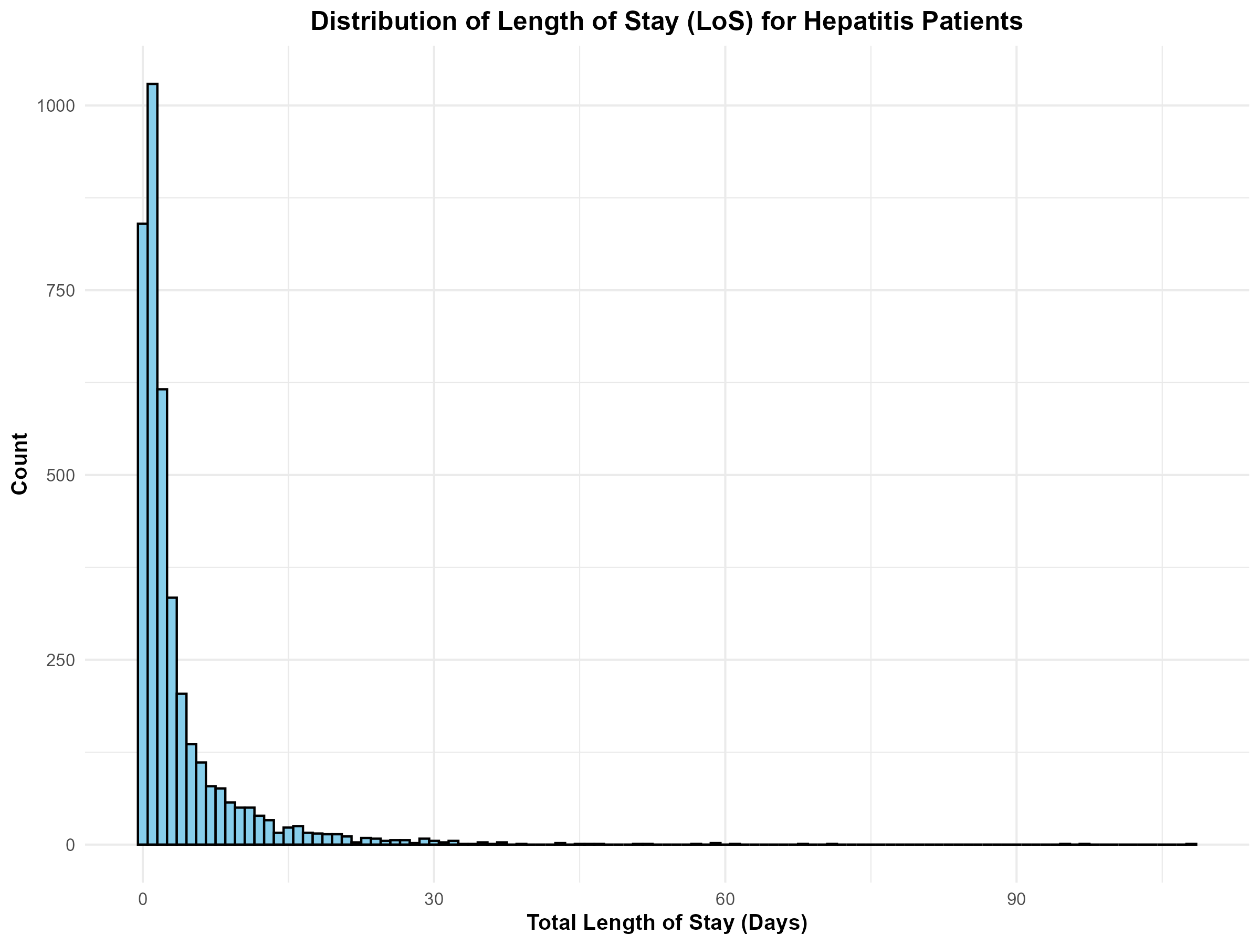

Sup Figure 1. Distribution of total LoS for hepatitis patients.

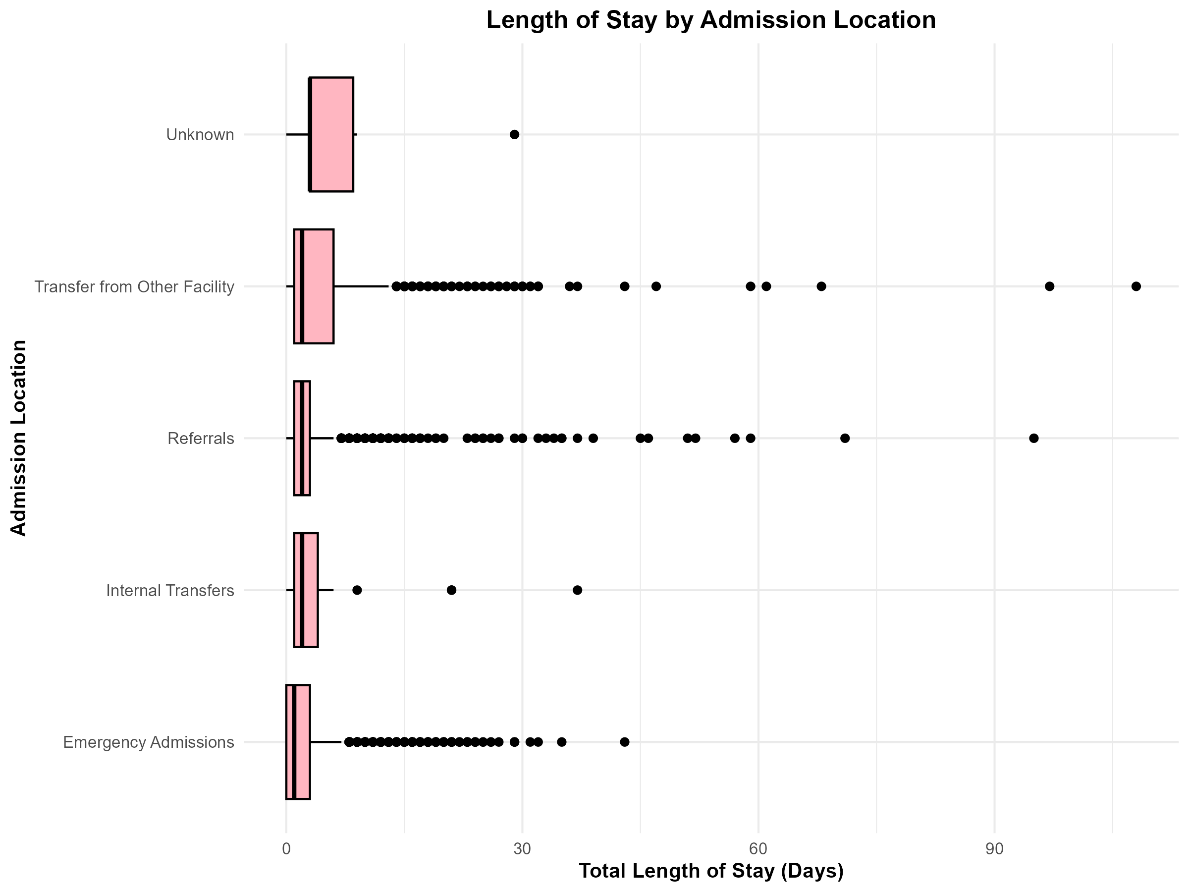

Sup Figure 2. Length of stay by admission location.

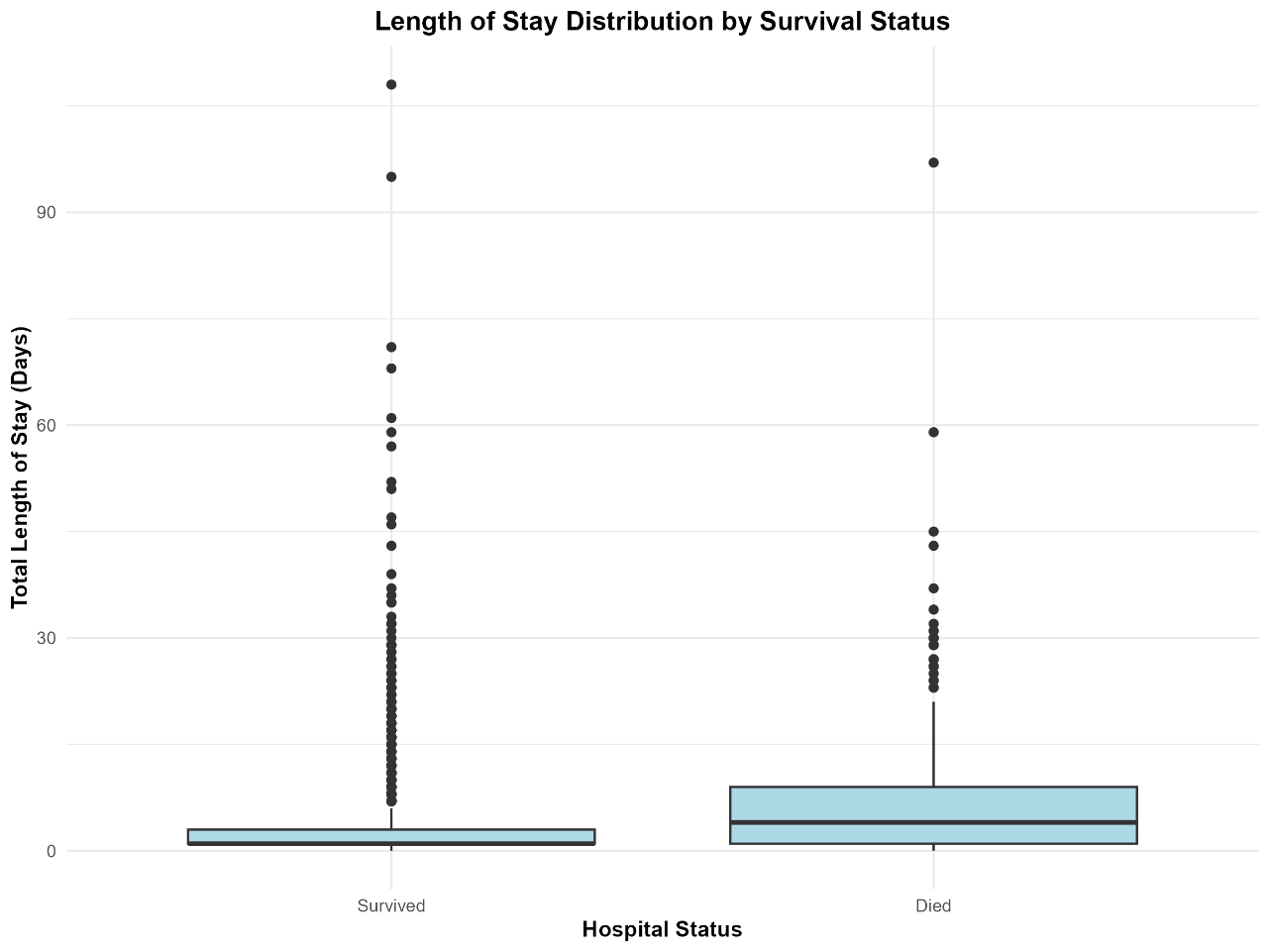

Sup Figure 3. Discharge outcome based on total LoS.

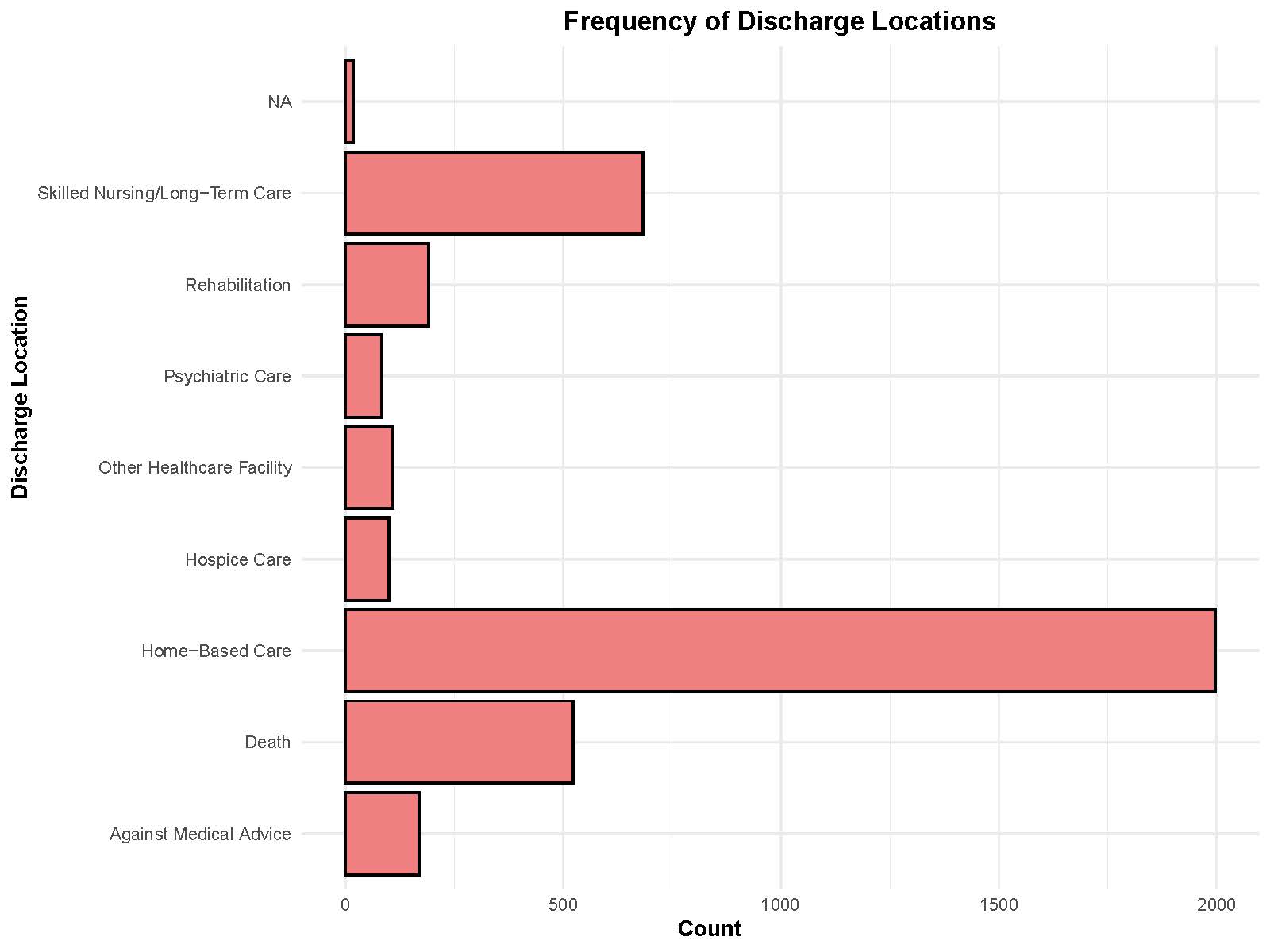

Sup Figure 4. Distribution of discharge locations among patients.

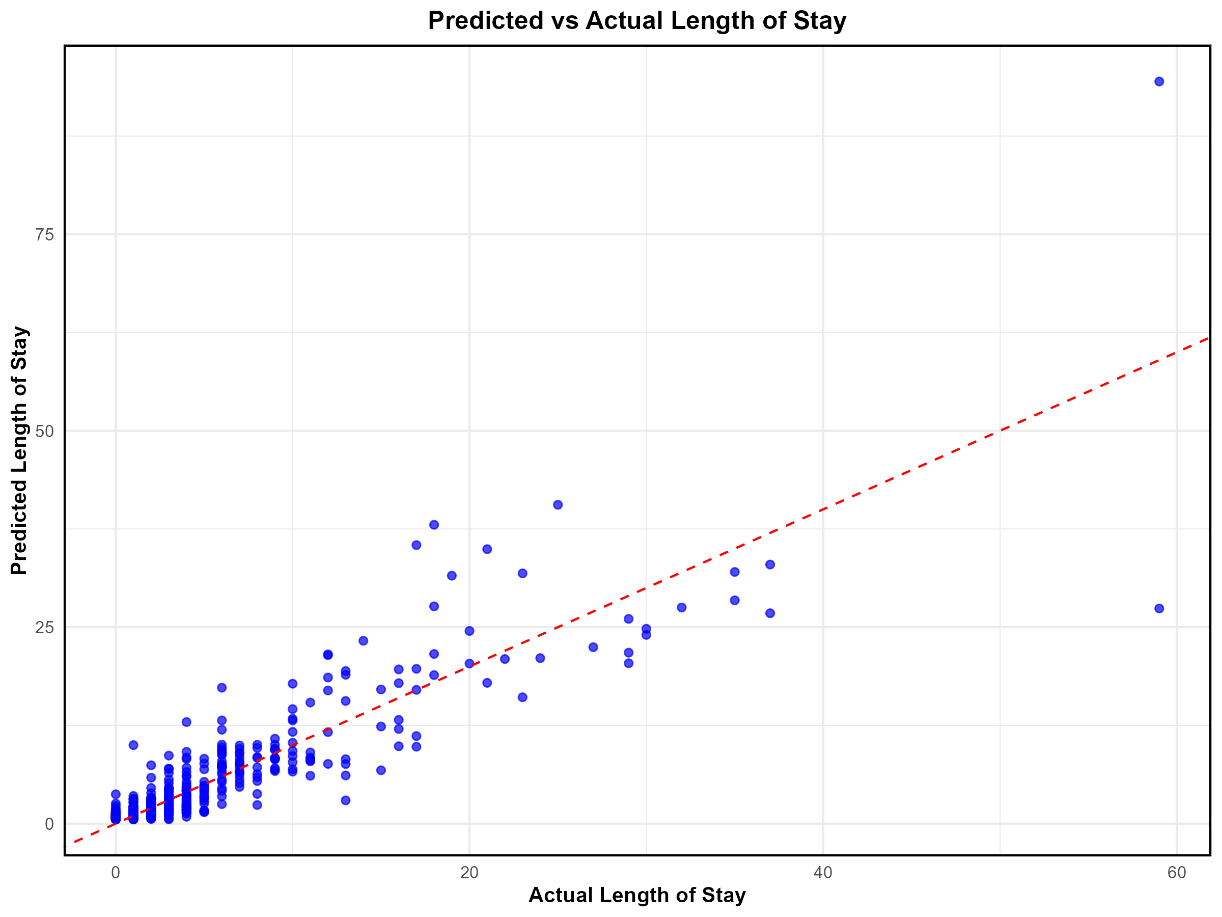

Sup Figure 5. Predicted vs. Actual Length of Stay for the Generalized Additive Model (GAM)

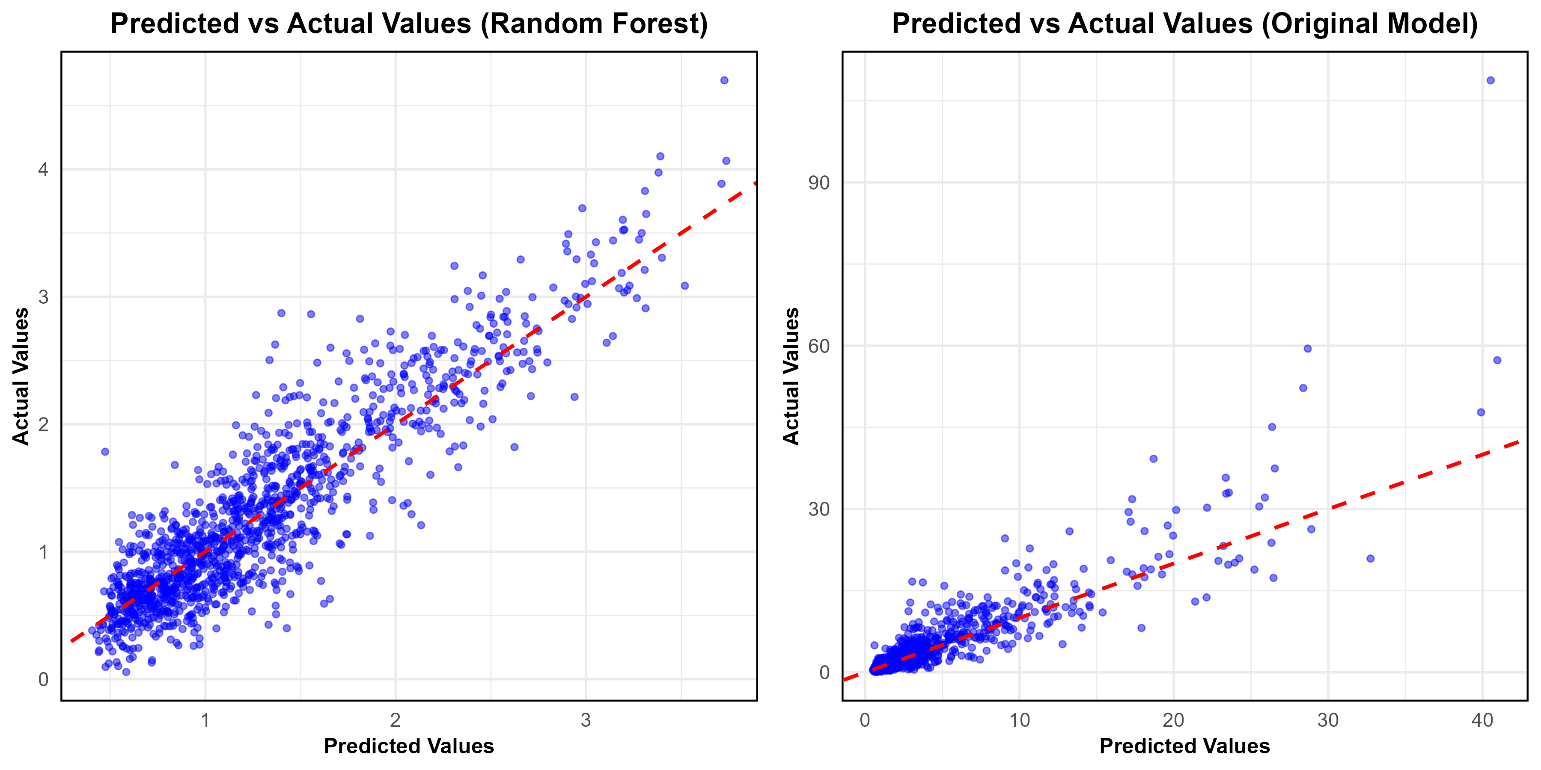

Sup Figure 6. Predicted vs. Actual Length of Stay for the Random Forest Regression Model: (Left) Log-Transformed Data and (Right) Original Data.

**Definitions**

1. **Root Mean Squared Error (RMSE)**: Measures the average magnitude of error between predicted and actual values, penalizing larger deviations more heavily, making it particularly sensitive to outliers
2. **R-squared (R²)**: Quantifies the proportion of variation in the dependent variable explained by the model, offering an overall measure of fit
3. **Mean Absolute Error (MAE)**: Calculates the average of the absolute errors, providing an intuitive measure of model accuracy by treating all errors equally
4. **Accuracy**: Reflects the percentage of correctly classified instances.
5. **Kappa**: Accounts for agreement by chance, providing a more robust evaluation of model performance
6. **Brier Score**: Measures the mean squared difference between predicted probabilities and the actual class, with lower scores indicating better calibration
7. **Area Under the Receiver Operating Characteristic Curve (ROC AUC):** Quantifies a model's ability to distinguish between classes, with values closer to 1 denoting superior performance.
8. **Sensitivity (also known as recall)**: Indicates the proportion of true positives correctly identified by the model.
9. **Specificity:** Measures the proportion of true negatives correctly identified.
10. **10-fold cross-validation**: This approach divides the dataset into 10 subsets (or folds). The model is trained on 9 folds and validated on the remaining fold in each iteration. This process is repeated 10 times, with each fold serving as the validation set once, and the average performance across all iterations is reported.

**R Packages**

Data processing was performed using the *tidyverse(41)*, while missing data were imputed with *mice* (42), and class imbalance was addressed using *DMwR (26,44)*. Model development and evaluation were facilitated by *MachineShop* (43), enabling a streamlined approach to implementing and comparing various predictive models. Statistical modeling was conducted using *MASS* (45), which supported the fitting of Negative Binomial regression models, and *mgcv* (46), which was utilized for constructing GAMs to capture nonlinear relationships. Random Forest modeling was implemented using the *randomForest* package (48). Data visualizations were created with *ggplot2* (47), while descriptive statistics and baseline characteristics were summarized using *tableone* (49).
